# Dual Antiplatelet Therapy (DAPT) in Patients with NSTEMI Undergoing CABG

**DOI:** 10.1101/2025.02.13.25322259

**Authors:** Agam Bansal, Grant W. Reed, Tom Wang, Jacqueline Tamis-Holland, Wael A. Jaber, Samir R. Kapadia, Venu Menon

**Author notes:** Corresponding author Venu Menon, M.D. FACC, FAHA, FESC Director Cardiac Intensive Care Unit Director Cardiovascular Fellowship Professor of Medicine Heart and Vascular Institute, Cleveland Clinic, Cleveland, Ohio.

## Abstract

**Background:** Dual antiplatelet therapy (DAPT), combining acetylsalicylic acid (aspirin) and a P2Y12 inhibitor, is recommended for patients with non-ST segment elevation myocardial infarction (NSTEMI) undergoing coronary artery bypass grafting (CABG). However, real-world adoption and effectiveness of DAPT in this context has not been adequately evaluated.

**Methods:** We conducted a retrospective propensity-matched cohort analysis using data from the TriNetX research network, which includes electronic health records from over 70 healthcare organizations across the United States. The study included patients aged >18 years diagnosed with NSTEMI who underwent CABG within one month of presentation from January 2015 to January 2024. Outcomes for patients on DAPT were compared with those on aspirin monotherapy using standardized mean differences, risk ratios, and Cox proportional hazard models for survival analysis.

**Results:** From a cohort of 21,092 NSTEMI patients eligible for DAPT post-CABG, 55.28% received DAPT predominantly consisting of aspirin and clopidogrel. After propensity score matching, DAPT was associated with significantly reduced all-cause mortality at 1-year (8.1% vs 5.5%, OR: 1.52) and 5-year (14.4% vs 10.6%, OR: 1.41) follow-ups compared to aspirin monotherapy. There were no significant differences in rates of major bleeding, ischemic strokes, or repeat revascularization between the two groups.

**Conclusion:** The underutilization of DAPT in real-world settings, despite guideline recommendations, reflects a potential gap between clinical practice and evidence-based guidelines. Our findings support the effectiveness of DAPT in reducing mortality without increasing major bleeding risks, underscoring the need for more widespread adoption and potentially more robust clinical trials to confirm these observational findings.

## Introduction

An early invasive strategy is the treatment of choice in high-risk patients presenting with non-ST segment elevation myocardial infarction (NSTEMI) (1). While the dominant modality of revascularization following NSTEMI is percutaneous coronary intervention (PCI), approximately 10-16% of the subjects enrolled in the randomized trials (2,3) were referred for initial coronary artery bypass graft (CABG) surgery. Current guidelines (1,4) provide a Class I recommendation for continued dual antiplatelet therapy (DAPT) with acetylsalicylic acid (aspirin) and a P2Y_12_ inhibitor in patients with NSTEMI undergoing CABG with an endorsement to utilize a more potent P2Y_12_ inhibitor (ticagrelor) and to continue DAPT for a duration of 12 months. These recommendations are based on expert consensus opinion (level C) and supported in part by post randomization observations in patients undergoing CABG in the early trials exploring the utilization and composition of DAPT (2,3,5) following NSTEMI. This recommendation is also supported by meta-analysis (6,7), small, randomized trials (8,9) with surrogate end points such as vein graft failure and data from observational registries. There are no adequately powered randomized clinical trials supporting the current post CABG DAPT recommendations. The adoption of this guideline recommendation by clinicians in the United States remains uncertain. Given this gap in evidence, we sought to evaluate the utilization of DAPT after CABG and associated clinical outcomes in a contemporary real-world registry.

## Methodology

### Data Source

The data analyzed in this study were obtained from the TriNetX research network, which contains data from electronic health records of more than 90 million patients from over 70 health care organizations, primarily in the United States. TriNetX provides data including demographics, diagnostic and procedural information, and standard measurements (including vital signs, laboratory results, and medications) using standardized coding systems (International Classification of Diseases-10th Revision-Clinical Modification [ICD-10-CM] and Current Procedural Terminology codes for diagnoses and procedures, Logical Observation Identifiers Names and Codes for vital signs and laboratory values, and RxNorm for medications).

### Study Population and Design

We performed a retrospective propensity-matched cohort analysis on patients aged ≥ 18 years with non-ST-segment elevation myocardial infarction (NSTEMI) who underwent CABG within 1 month of presentation from January 1^st^, 2015, until 7^th^ January 2024. NSTEMI was defined using the ICD-10 codes and performance of CABG was identified using the Current Procedural Terminology (CPT) codes. The prescription of different antiplatelet therapy regimens including acetylsalicylic acid (aspirin) and P2Y_12_ inhibitors (clopidogrel/ticagrelor/prasugrel) were ascertained within 2 weeks of CABG. Patients on anticoagulation with warfarin/apixaban/rivaroxaban/dabigatran were excluded from the analysis.

### Study Outcomes

Utilization of different antiplatelet therapies (aspirin monotherapy or DAPT) were determined in patients with NSTEMI after CABG. We compared 1- and 5-year outcomes between patients on aspirin monotherapy versus DAPT. The primary efficacy outcome assessed was all-cause mortality. Secondary outcomes included major bleeding requiring blood transfusion, ischemic/hemorrhagic stroke, need for repeat revascularization (PCI or redo CABG) and performance of venous graft percutaneous revascularization. These endpoints were ascertained using the ICD-10 and CPT codes.

### Statistical Analysis

Continuous variables are represented as mean ± SD and were compared between the groups using independent-samples Student’s t-tests. Categorical variables are reported as counts (percentages) and were compared between the groups using the chi-square test. Covariates were matched extensively by 1:1 propensity score matching (PSM) using the greedy nearest-neighbor algorithm with a caliper of 0.1 pooled standardized mean difference (SMD). The standardized mean difference is a quantitative method used to represent the difference between the means of two groups in terms of SD units to assess the balance in measured variables in the sample weighted by the inverse probability of treatment. Any characteristic with a between cohorts lower than 0.1 was considered well matched. The measures of association included risk differences, risk ratios, and odds ratios (ORs) on the matched population for primary and secondary outcomes. Survival analyses were performed for outcomes by plotting Kaplan-Meier curves with log-rank tests. Additionally, Cox proportional hazard models were used to calculate the hazard ratios to compare the two groups. Death was treated as a censoring event. Further, the temporal trend analysis was performed using the Cochrane Armitage trend test. Statistical significance was set at a 2-sided p-value of P<0.05. Statistical analyses were completed using the TriNetX online platform using R for statistical computing.

### Study Oversight

Data were analyzed and interpreted by the authors. All authors reviewed the manuscript and affirmed the accuracy and completeness of the data. Institutional review board (IRB) approval was exempt given that aggregate deidentified data were used from the research network database. These study findings are reported per the Strengthening the Reporting of Observational Studies in Epidemiology (STROBE) guidelines for cohort studies.

## Results

### Study Population

A total of 523,624 patients were hospitalized with NSTEMI from January 1^st^, 2015, until January 7^th^, 2024. Overall, 5.08% (26,606/523624) underwent CABG within 1 month of hospitalization. After excluding patients on anticoagulation, a total of 21,092 patients with NSTEMI were identified to be potential candidates for DAPT after CABG. [Figure 1]. The mean age of the cohort was 69 ± 11 years, 70% were males, and 75% were Caucasians.

**Figure 1:**
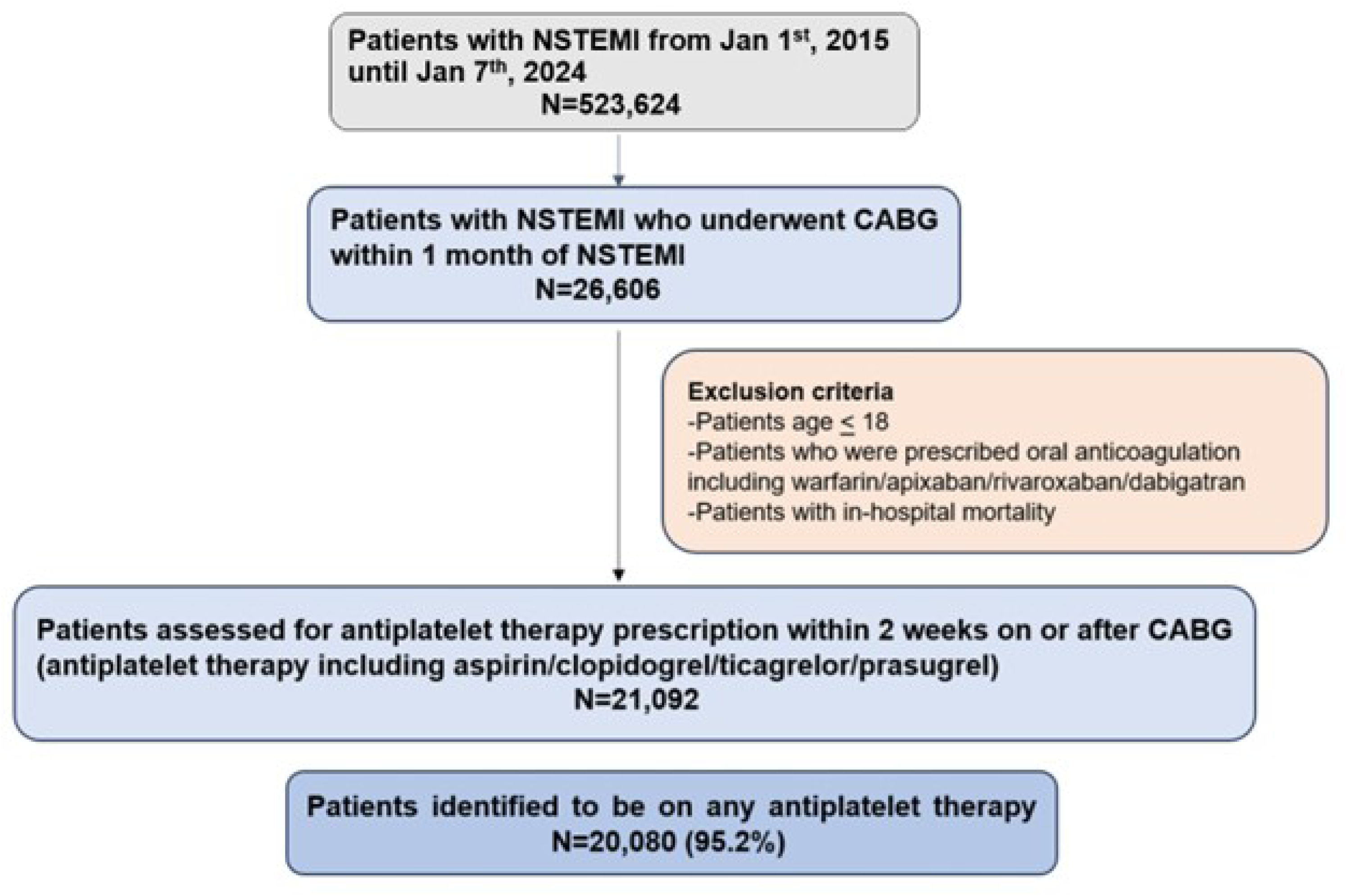
Study methodology

### Utilization of Antiplatelet Therapy

Of the 21,092 patients, the data regarding antiplatelet therapy regimen prescribed within 2 weeks on or after CABG was available for 95.2% (N=20,080) individuals. DAPT (aspirin and clopidogrel/ticagrelor/prasugrel) was utilized in 55.28% patients (11,101/20,080) [Central illustration]. The most common DAPT regimen was the combination of aspirin and clopidogrel (97.06%, N=10,775/11,101). Aspirin monotherapy was utilized in 42.21% (N=8,477/20,080). [Figure 2]. Amongst patients with NSTEMI undergoing CABG, the utilization of DAPT increased significantly over the years (34.20% in 2015 and 56.3% in 2023), p<0.001. [Figure 3].

**Figure 2:**
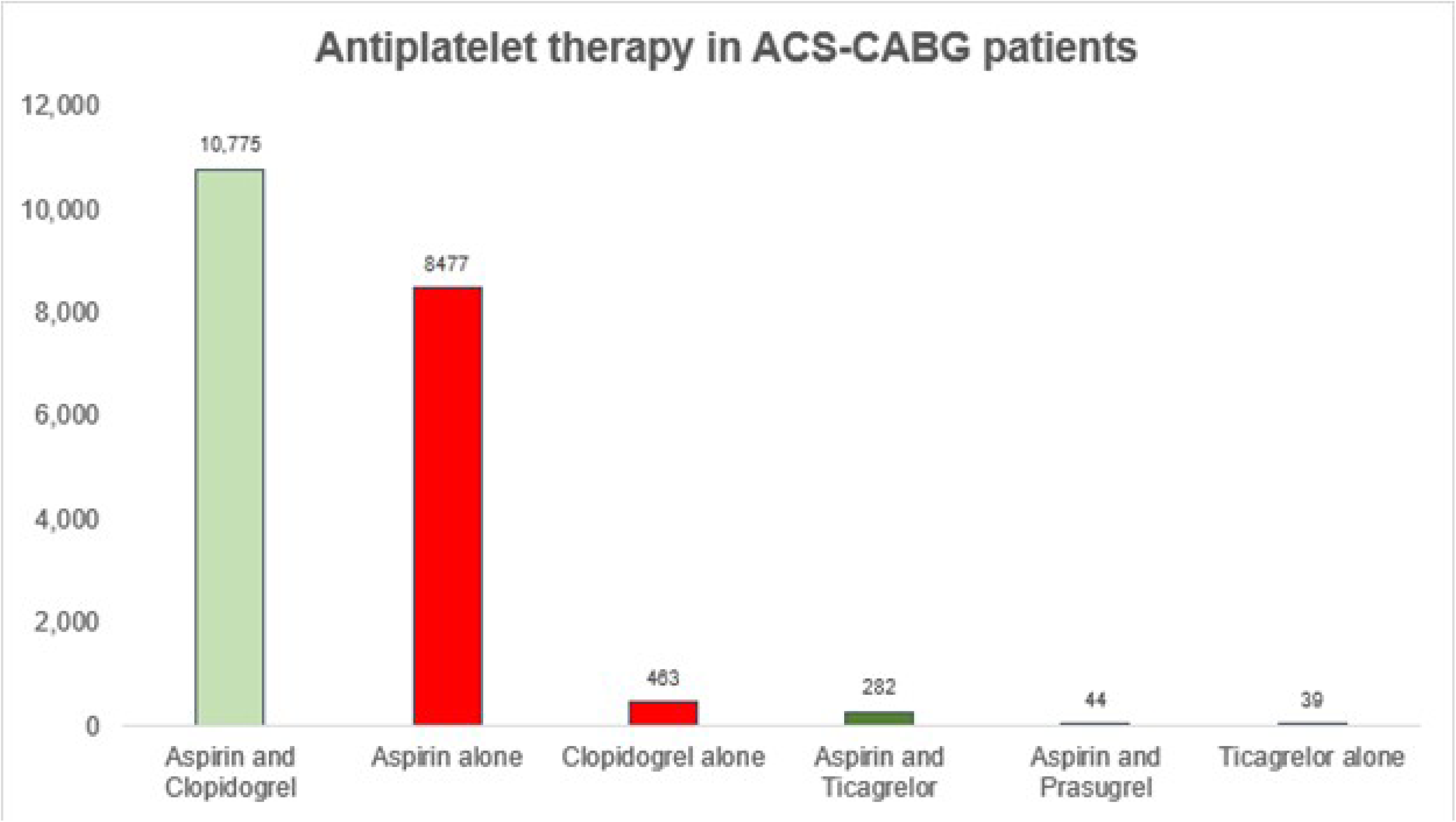
Utilization of antiplatelet therapy in ACS-CABG patients.

**Figure 3:**
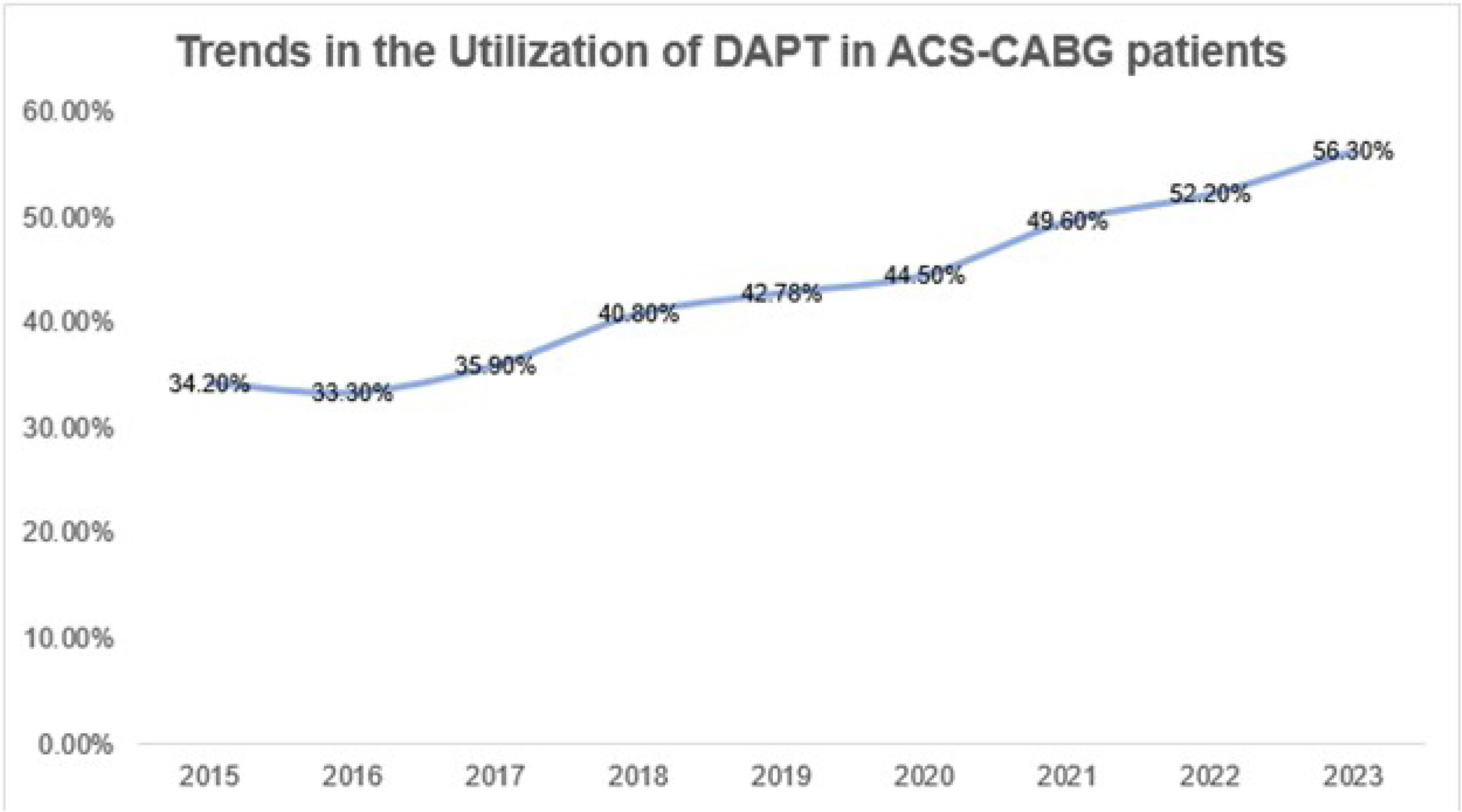
Trends in the Utilization of DAPT in ACS-CABG patients

### Aspirin monotherapy versus DAPT

#### Patient characteristics

Baseline characteristics of the patients undergoing CABG based on utilization of DAPT before and after PSM, are presented in Table 1. As compared to aspirin monotherapy, patients on DAPT were younger, more likely to be Caucasian, and had higher prevalence of hyperlipidemia, smoking, prior myocardial infarction, prior percutaneous coronary intervention (PCI), prior stroke, peripheral vascular disease, and COPD. Patients with chronic kidney disease (CKD) or end stage renal disease (ESRD) were more likely to be on aspirin monotherapy. The DAPT group was also more likely to be prescribed statins, beta-blockers, and angiotensin receptor blockers. The left ventricular ejection fraction was similar in both groups. After PSM, baseline characteristics in the two groups were similar and no residual imbalances were noted (standard difference: <0.1 for all covariates).

**Table 1:** Characteristics of ACS-CABG Patients who were Prescribed DAPT (ASA+ Clopidogrel/Ticagrelor/Prasugrel) versus ASA monotherapy

|  | Aspirin<br>(N=8477) | DAPT<br>(N=11,101) | p-value | Aspirin<br>(N=7699) | DAPT<br>(N=7699) | p-value |
| --- | --- | --- | --- | --- | --- | --- |
|  | Before propensity score matching |  |  | After propensity score matching |  |  |
| Characteristics |  |  |  |  |  |  |
| Age at index admission | 64.8 ± 10.6 | 63.9 ± 10.4 | <0.001 | 64.6 ± 10.6 | 64.5 ± 10.4 | 0.805 |
| Females | 25.0% | 25.49% | 0.446 | 25.0% | 24.8% | 0.744 |
| White | 72.0% | 74.9% | <0.001 | 72.0% | 72.1% | 0.838 |
| Hypertension | 81.0% | 81.9% | 0.131 | 81.4% | 81.5% | 0.915 |
| Diabetes mellitus | 57.4% | 57.3% | 0.831 | 57.4% | 56.7% | 0.420 |
| Hyperlipidemia | 82.7% | 85.6% | <0.001 | 83.4% | 83.3% | 0.771 |
| Smoking | 29.1% | 35.0% | <0.001 | 29.1% | 29.5% | 0.623 |
| Chronic kidney disease | 32.1% | 27.4% | <0.001 | 31.1% | 30.6% | 0.407 |
| ESRD | 10.2% | 7.3% | <0.001 | 9.4% | 9.1% | 0.585 |
| Peripheral vascular disease | 15.1% | 17.8% | <0.001 | 15.2% | 15.7% | 0.321 |
| Prior MI | 23.8% | 29.1% | <0.001 | 24.5% | 24.7% | 0.787 |
| Prior PCI | 15.4% | 21.8% | <0.001 | 16.3% | 16.1% | 0.821 |
| COPD | 16.6% | 18.7% | <0.001 | 16.7% | 16.8% | 0.929 |
| Prior stroke | 7.7% | 9.5% | <0.001 | 8.1% | 8.3% | 0.671 |
| Medications |  |  |  |  |  |  |
| Statins | 94.5% | 97.0% | <0.001 | 95.9% | 95.8% | 0.867 |
| Beta blockers | 87.2% | 93.4% | <0.001 | 91.2% | 90.8% | 0.561 |
| ACE inhibitors | 47.0% | 46.8% | 0.805 | 47.5% | 47.8% | 0.678 |
| ARBs | 23.6% | 26.5% | <0.001 | 24.5% | 24.5% | 0.908 |
| Laboratory values |  |  |  |  |  |  |
| Serum creatinine | 1.31 ± 1.46 | 1.22 ± 1.55 | <0.001 | 1.3 ± 1.45 | 1.27 ± 1.31 | 0.167 |
| LV Ejection fraction, % | 50.2 ± 14.3 | 49.5 ± 15.2 | 0.099 | 50.3 ± 14.3 | 49.5 ± 15.4 | 0.184 |

ESRD: end stage renal disease; PCI: percutaneous coronary intervention; MI: myocardial infarction; COPD: chronic obstructive pulmonary disease; ACE: angiotensin converting enzyme; ARBs: angiotensin receptor blockers; LV: left ventricular.

### Clinical Outcomes

#### 1-year outcomes

After PSM, the primary outcome of all-cause mortality was significant higher in the aspirin monotherapy group (8.1% vs 5.5%, odds ratio (OR): 1.52 (1.33-1.73), p<0.001) [Table 2]. The probability of survival at the end of 1-year was significantly higher in the DAPT group (93.6% vs 91.1%; hazard ratio (HR): 1.45 (1.28-1.64), log-rank test: X^2^ = 34.2, df=1, p<0.001) [Figure 4a, central illustration]. There were no differences in rates of ischemic stroke (1.3% vs 1.4%; aOR (0.95 (0.71-1.26), p=0.716)), repeat revascularization (PCI or redo CABG) (2.9% vs 3.2%, aOR: 0.92 (0.76-1.11), p=0.366)), and venous graft PCI interventions (0.88% vs 0.84%, aOR: 1.05 (0.74-1.49), p=0.789)) between the two groups. [Table 2].

**Table 2:** 1-year outcomes of DAPT vs ASA monotherapy in ACS-CABG patients

|  | Aspirin<br>(N=8477) | DAPT<br>(N=11,101) | p-value | Aspirin<br>(N=7699) | DAPT<br>(N=7699) | p-value |
| --- | --- | --- | --- | --- | --- | --- |
|  | Before propensity score matching |  |  | After propensity score matching |  |  |
| 1-year Outcomes |  |  |  |  |  |  |
| All-cause mortality | 8.0% | 5.2% | 1.58 (1.41-1.77),<br>p<0.001 | 8.1% | 5.5% | 1.52 (1.33-1.73), p<0.001 |
| Need for blood transfusion | 5.4% | 5.7% | 0.96 (0.85-1.08), p=0.554 | 5.8% | 5.9% | 0.99 (0.86-1.13), p=0.889 |
| Stroke | 1.4% | 1.6% | 0.83 (0.66-1.06), p=0.140 | 1.3% | 1.4% | 0.95 (0.71-1.26), p=0.716 |
| Ischemic stroke | 1.3% | 1.5% | 0.80 (0.62-1.03), p=0.100 | 1.1% | 1.2% | 0.90 (0.66-1.20), p=0.443 |
| Hemorrhagic stroke | 0.16% | 0.17% | 0.91 (0.45-1.86), p=0.815 | 0.17% | 0.15% | 1.30 (0.57-2.96), p=0.531 |
| Repeat revascularization (PCI or redo CABG) | 3.0% | 3.4% | 0.94 (0.84-1.22), p=0.489 | 2.9% | 3.2% | 0.92 (0.76-1.11), p=0.366 |
| Venous graft percutaneous revascularization | 0.85% | 0.87% | 0.97 (0.72-1.32), p=0.858 | 0.88% | 0.84% | 1.05 (0.74-1.49), p=0.789 |

**Figure 4.**
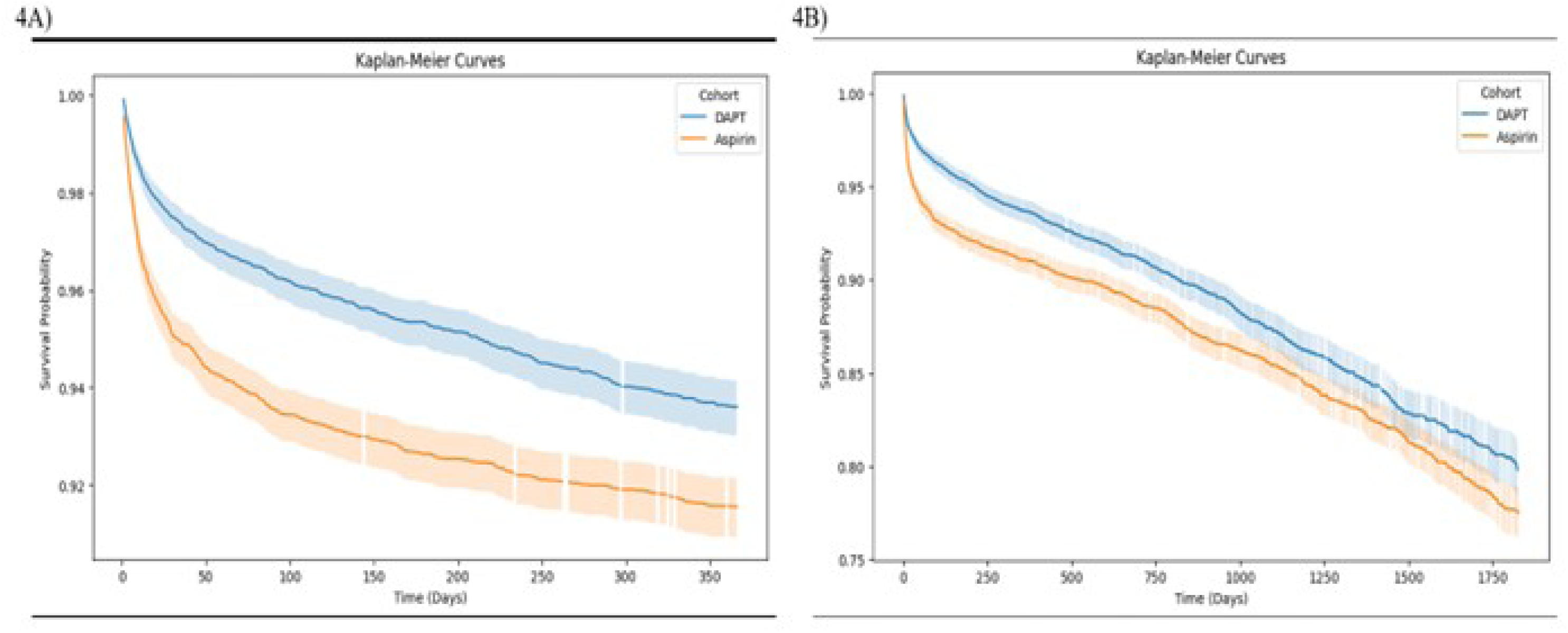
**4a: Kaplan Meier Curve for All-cause Mortality at 1-year.** The probability of survival at the end of 1-year was significantly higher in the DAPT group (93.6% vs 91.1%; hazard ratio (HR): 1.45 (1.28-1.64), log-rank test: X^2^ = 34.2, df=1, p<0.001) **4b: Kaplan Meier Curve for All-cause Mortality at 5-years:** The probability of survival at the end of 5-year was significantly higher in the DAPT group (79.4% vs 76.4%; hazard ratio (HR): 1.25 (1.11-1.36), log-rank test: X^2^ = 14.39, df=1, p<0.001)

With regards to safety, there was no significant difference in the need for blood transfusion between the two groups (5.8% vs 5.9%, OR: 0.99 (0.86-1.13), p=0.889) [Table 2]. There were also no differences in event free survival for blood transfusion between the two groups (HR: 93.8% vs 93.5%), log-rank test: X^2^ = 0.054, df=1, p=0.816) [Figure 5] and rates of hemorrhagic stroke (0.17% vs 0.15%; aOR (1.30 (0.57-2.96), p=0.531)) were similar.

#### *5-year* outcomes

After PSM, the primary outcome of all-cause mortality was significant higher in the aspirin monotherapy group (14.4% vs 10.6%, odds ratio (OR): 1.41 (1.28-1.59), p<0.001) [Table 3]. Furthermore, the probability of survival at the end of 5-year was significantly higher in the DAPT group (79.4% vs 76.4%; hazard ratio (HR): 1.25 (1.11-1.36), log-rank test: X^2^ = 14.39, df=1, p<0.001) [Figure 4b, central illustration].

**Table 3:**
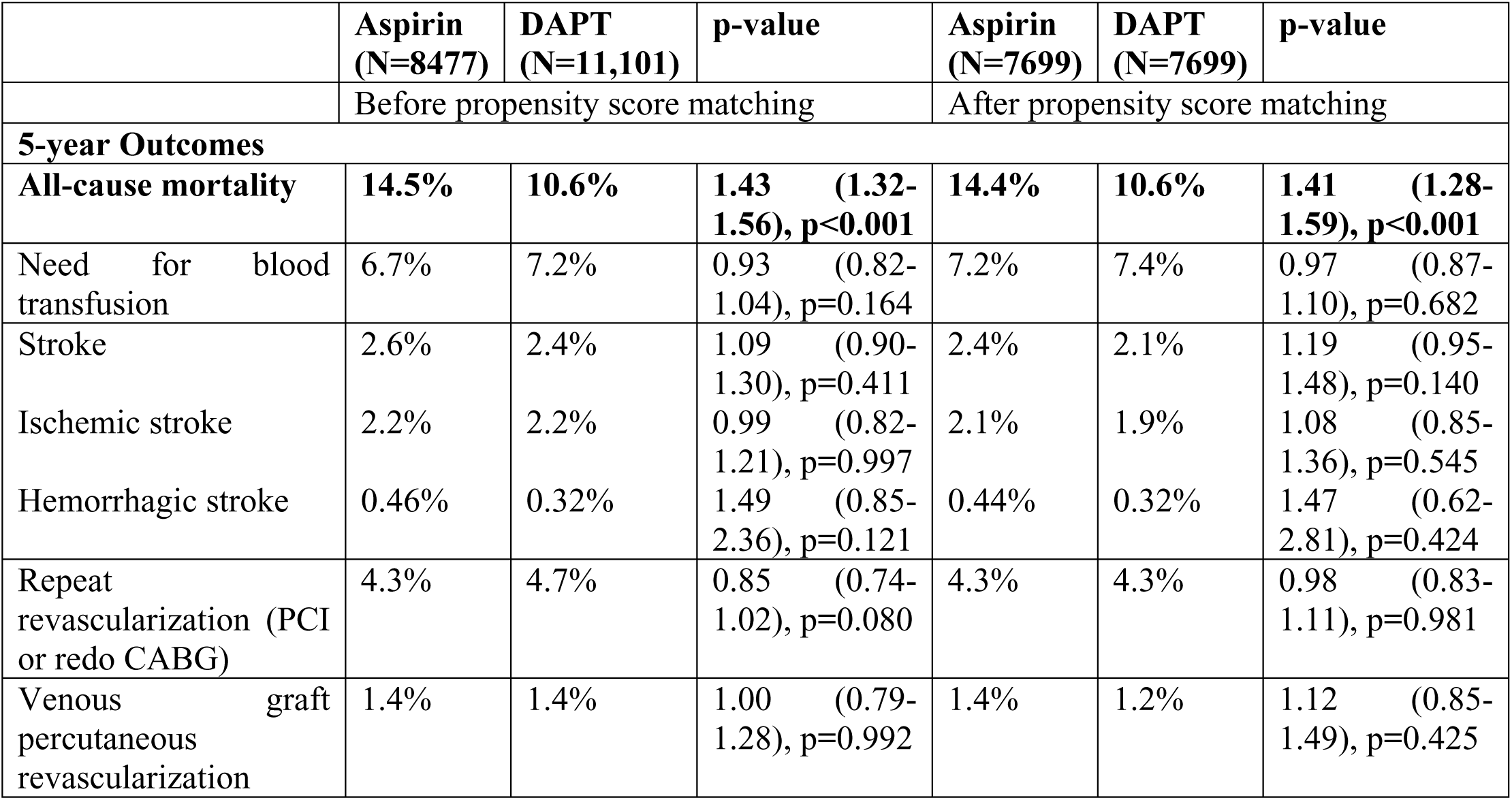
5-year outcomes of DAPT vs ASA monotherapy in ACS-CABG patients

There was no significant difference in the need for blood transfusion between the two groups (7.2% vs 7.4%, OR: 0.97 (0.87-1.10), p=0.682) [Table 3]. There was no difference in event free survival for blood transfusion between the two groups (HR: 0.93 (0.83-1.05), log-rank test: X^2^ = 1.51, df=1, p=0.221)

There were no differences in ischemic stroke (2.1% vs 1.9% (aOR: 1.19 (0.95-1.49), p=0.140)), hemorrhagic stroke (0.44% vs 0.32%, aOR: 1.47 (0.62-2.81), p=0.424)), repeat revascularization (PCI or redo CABG) (4.3% vs 4.3%, aOR: 0.98 (0.83-1.11), p=0.981)), and venous graft PCI interventions (1.4% vs 1.2%, aOR: 1.12 (0.85-1.49), p=0.425)) between the two cohorts. [Table 3].

## Discussion

We performed a large, retrospective, observational study to evaluate the utilization and outcomes associated with DAPT exposure following CABG for NSTEMI in a contemporary US (United States) registry. The salient findings of our study are: 1) the utilization of DAPT following NSTEMI-CABG is limited but increasing over time; 2) clopidogrel is the predominant P2Y_12_ inhibitor utilized in this setting; 3) DAPT in comparison with ASA monotherapy is associated with a significantly reduced all-cause mortality; and 4) there are no significant differences in hemorrhagic stroke, need for blood transfusions, and repeat revascularization with DAPT utilization.

Current American College of Cardiology/American Heart Association (ACC/AHA) (4) and European Society of Cardiology (ESC) (1) guidelines recommend DAPT continuation with a potent P2Y_12_ inhibitor in patients with ACS undergoing CABG for a duration of 12 months (Class I recommendation, level of evidence C). Despite this recommendation supporting DAPT utilization in a NSTEMI-CABG population, adoption of this guidance in real world practice appears low with only about a half of the post CABG patients receiving DAPT. This likely reflects a lack of clinician confidence in these post hoc observations and is consistent with that noted in prior studies and registries. In the Danish (10) and Swedish national registries (11), around 30% of the ACS-CABG patients were treated with DAPT and similar rates of DAPT utilization have been reported from Canada (12) and Korea (13). We did however observe a significant increase in the utilization of DAPT from the years 2016 (33.3%) to 2023 (56.3%) likely influenced by the consistent support for DAPT in ACS-CABG patients in ACC/AHA guidelines in 2016 (14) and ESC guidelines in 2017 (15) that have remained consistent in several subsequent guidelines (16, 17).

With respect to the choice of P2Y_12_ inhibitors, ticagrelor or prasugrel are recommended over clopidogrel if the patient is not at a high risk of bleeding. This guidance is supported by the findings from subgroup of CABG patients in the Platelet Inhibitor and Patient Outcomes (PLATO) trial (18) and the TRITON TIMI 38 study (19) demonstrating superior clinical outcomes with ticagrelor and prasugrel over clopidogrel respectively. In our study, clopidogrel was overwhelming utilized as the part of DAPT regimen (97%). These findings are also consistent with large observational studies from Denmark (10) and Korea (13) whereas ticagrelor was the predominant P2Y_12_ inhibitor in the Swedish Cardiac Registry (11).

Patients undergoing CABG prescribed DAPT were more likely to have risk factors representative of increased ischemic risk such as prior myocardial infarction or PCI, heart failure, smoking, or peripheral vascular disease, whereas DAPT was less likely to be dispensed in post CABG patients with renal dysfunction, potentially because of the increased bleeding risk. The decision to utilize clopidogrel exclusively in this US registry may be driven by a generalized concern for the risk of bleeding rather than guided by assessment of individual bleeding/ischemia risk. Although the benefits of ticagrelor and prasugrel over clopidogrel are amplified in patients with diabetes mellitus; the utilization of DAPT in patient with and without diabetes were similar with minimal use of potent P2Y12 inhibitors.

NSTEMI-CABG patients on DAPT in the current registry had a substantially lower observed all-cause mortality rate without a significant increase in bleeding requiring blood transfusion or hemorrhagic stroke. This favorable finding with DAPT persisted despite propensity matching. Similar clinical benefits have been noted in some but not all prior studies. In a subgroup analysis of patients undergoing CABG in the CURE trial (20), randomization to aspirin plus clopidogrel was shown to reduce the composite of cardiac death, myocardial infarction, or stroke without a significant increase in life-threatening bleeding; however, benefits were mainly seen prior to revascularization. In PLATO, clinical outcomes following the CABG were significantly improved in subjects randomized to ticagrelor over clopidogrel with a pronounced reduction in observed CV (4.1 vs. 7.9%, p=0.0092; HR 0.52; 95% CI (0.32-0.85) and all-cause mortality (4.7 vs. 9.7%, p=0.0018; HR 0.49; 95% CI (0.32-0.77). Data from existing large observational studies (10, 11, 13) have yielded discrepant results for the benefit of DAPT in ACS-CABG patients. While the studies from Denmark (10) and Korea (13) have demonstrated a significant reduction in mortality and recurrent myocardial infarction with DAPT, there were no significant differences in the risk of major adverse cardiovascular events between the DAPT (ASA and ticagrelor) versus aspirin group, in a large 6558 patient Swedish Registry with increased risk for major bleeding (11). The TACSI trial is a prospective, multicenter, open-label-registry-based randomized trial that is testing the hypothesis that 1-year treatment with DAPT consisting of ticagrelor and aspirin is superior to aspirin alone after isolated CABG in ACS patients. In the setting of conflicting results from the observational studies, findings of the ongoing TACSI trial (NCT03560310) (21) will be highly valuable.

In our study, around 5% of NSTEMI patients underwent CABG within 1 month of NSTEMI. These rates are lower than noted in the RCTs (2,3) (CURE-16% and PLATO-10%). In the Medicare dataset (22), there was noted to be a decline in the utilization of CABG for AMI from 14.4% in 1995 to 10.2% in 2014 with a concurrent increase in PCI from 18.8% to 43.3%. Similarly, in the national inpatient dataset (23), there was a significant decline in the utilization of CABG from 10.5% in 2000 to 8.7% in 2017. There are several reasons that could account for reduced rates of CABG in NSTEMI population in our study. The success of catheter-based-management and improvement in medical treatment has reduced the need for urgent/emergent CABG in patients presenting with AMI (24). It is also plausible that the threshold to perform revascularization with CABG has increased in clinical practice. Further, one of the limitations of our dataset is that it only captures outcomes if a patient remains in the same or participating healthcare organizations (HCOs), hence it possible that we may be underestimating CABG surgeries performed. Alternatively, in the troponin era, the proportion of patients with myocardial injury erroneously coded as having a NSTEMI may also contribute to these findings.

To the best of our knowledge, this is the largest study to date to assess the utilization and long-term outcomes of DAPT in NSTEMI treated with CABG patients. However, our study has several limitations that must be acknowledged. First, this is an observational study with a risk for selection bias and residual confounding. While the degree and directionality of clinical outcome were similar before and after propensity matched analysis, there may be unmeasured factors impacting the decision to prescribe DAPT, and it is possible the “healthier” group of patients were treated with DAPT. Second, the outcomes analyzed in our study were described utilizing the ICD-10 and CPT codes and were not centrally adjudicated, hence there is an inherent limitation related to miscoding. Using the ICD-10 diagnosis codes, we were unable to specifically ascertain the Bleeding Academic Research Consortium (BARC) type bleeding events or life-threatening bleeding, hence we used the need for blood transfusion as a surrogate but objective metric for that endpoint. Importantly, we were unable to determine the incidence of recurrent myocardial infarction using the ICD-10 diagnoses codes. Although the ICD-10 code-I22 depicts subsequent STEMI or NSTEMI, it has been shown to be under-representative of the true incidence of recurrent myocardial infarction. Third, using the database we were unable to specifically report on cardiovascular mortality. Further, the reason for reduced all-cause mortality in the DAPT cohort could not be determined. It is possible that DAPT leads to a decrease in incidence of recurrent MI, however that is purely speculative. Fourth, around 4.8% of the individuals did not have information on any antiplatelet therapy within 2 weeks of CABG. This could either be because they were not prescribed any antiplatelet therapy or because of missing information. Fifth, we were unable to determine the duration of DAPT prescription and its impact on NSTEMI-CABG patients. Sixth, given a small number of patients in the ticagrelor or prasugrel cohort, we were unable to compare the outcomes amongst different P2Y_12_ inhibitors. Seventh, the TriNetX database only captures outcomes if a patient remains in the same or participating healthcare organization, and thus there is a likelihood of missing some clinical events. Given a very small number of patients being prescribed ticagrelor or prasugrel as a part of DAPT, we were unable to compare the outcomes based on the type of P2Y_12_ inhibitor used.

Our study has important clinical implications. One may speculate that a lack of strong evidence, fear of bleeding complications, and a strong tradition of prescribing ASA monotherapy to all patients undergoing CABG might contribute to only about half of the individuals receiving DAPT (that too primarily with a less potent P2Y_12_ inhibitor). In the light of around 40% higher all-cause mortality with ASA monotherapy as compared with DAPT without a significant increase in the need for blood transfusions, our findings from the real-world cohort support the current guideline emphasis for the need for prescribing DAPT in NSTEMI-CABG patients. Sufficiently powered prospective randomized trials with relevant end points that compare outcomes with different antiplatelet strategies and duration after CABG are warranted.

## Conclusions

This large, nationwide, population-based study demonstrates that the adherence to guidelines recommendations for DAPT after CABG in ACS patients is limited. When utilized, the use of DAPT as compared with ASA monotherapy is associated with significantly reduced all-cause mortality without an increase in bleeding.

Conflicts of Interest/Disclosures: None

Funding Source: None

## Data Availability

NA

## Acknowledgements

None

---

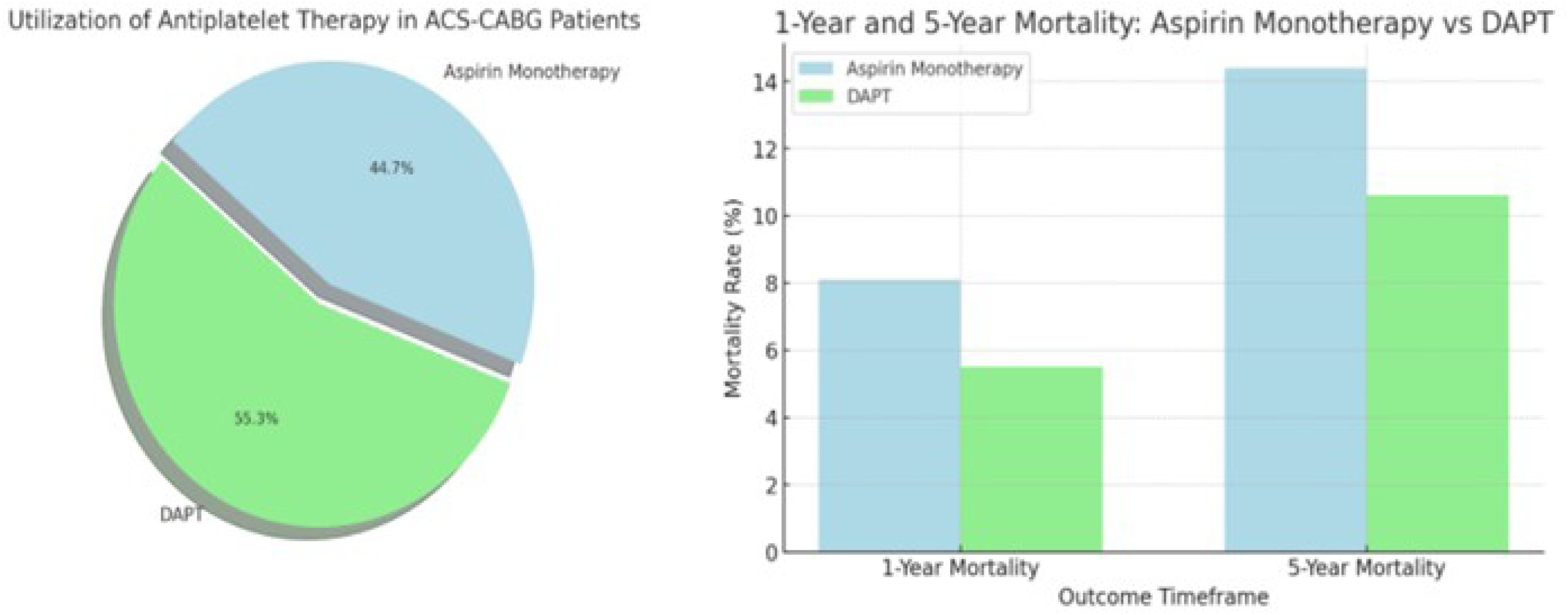
**Central Illustration:** Utilization and outcomes of DAPT in ACS-CABG patients

## Notes

### Competing Interest Statement

The authors have declared no competing interest.

### Funding Statement

There is no funding source

### Author Declarations

Protocol was exempt from institutional review board (IRB) approval given that aggregate de-identified data were used from research network database.

